# Exposure and risk factors for COVID-19 and the impact of staying home on Michigan residents

**DOI:** 10.1101/2020.08.25.20181800

**Authors:** Kuan-Han H. Wu, Whitney E. Hornsby, Bethany Klunder, Amelia Krause, Anisa Driscoll, John Kulka, Ryan Bickett-Hickok, Austin Fellows, Sarah Graham, Erin O. Kaleba, Salim S. Hayek, Xu Shi, Nadia R. Sutton, Nicholas Douville, Bhramar Mukherjee, Kenneth Jamerson, Chad M. Brummett, Cristen J. Willer

## Abstract

COVID-19 has had a substantial impact on clinical care and lifestyles globally. The State of Michigan reports over 80,000 positive COVID-19 tests between March 1, 2020 and July 29, 2020. We surveyed 8,047 Michigan Medicine biorepository participants in late June 2020. We found that 58% of COVID-19 cases reported no known exposure to family members or to someone outside the house diagnosed with COVID-19. A significantly higher rate of COVID-19 cases were employed as essential workers (45% vs 19%, p=3×10-11). COVID-19 cases reporting a fever were more likely to require hospitalization (categorized as severe; OR = 4.6 [95% CI: 1.7-13.0, p=0.004]) whereas respondents reporting rhinorrhea was less likely to require hospitalization (categorized as mild-to-moderate; OR = 0.16 [95% CI: 0.04-0.70, p=0.016]). African-Americans reported higher rates of being diagnosed with COVID-19 (OR = 4.0 [95% CI: 2.2-7.2, p=1×10-4]), as well as higher rates of exposure to family or someone outside the household diagnosed with COVID-19, an annual household income < $40,000, living in rental housing, and chronic diseases. During the Executive Order in Michigan, African Americans, women, and the lowest income group reported worsening health behaviors and higher overall concern for the potential detrimental effects of the pandemic. The higher risk of contracting COVID-19 observed among African Americans may be due to the increased rates of working as essential employees, lower socioeconomic status, and exposure to known positive cases. Continued efforts should focus on COVID-19 prevention and mitigation strategies, as well as address the inequality gaps that result in higher risks for both short-term and long-term health outcomes.

## INTRODUCTION

SARS-CoV-2 is a novel coronavirus that appears to have infected the first individual in Mainland China in December 2019. The virus has spread rapidly and globally, with documented cases in nearly every country, resulting in 16.8 million confirmed COVID-19 cases as of July 26, 2020 [1]. COVID-19 case numbers rapidly increased in the United States in March and April, particularly escalating in the states of New York, New Jersey, Connecticut, and Michigan. The rate of infection in Washtenaw County, Michigan, was 0.6% (2035 confirmed cases/367,601), as of July 26, 2020. Also on July 29, the State of Michigan reported 80,172 confirmed COVID-19 cases (0.8% of the state’s population) and 6,172 deaths, representing 2% of COVID-19 cases and 4% of deaths in the United States [2].

Limited information is available to fully explain why certain individuals appear to be at a higher risk. Based on the currently available data, older adults, especially those 65 years of age and older, are at the highest risk for hospitalization, intensive care, ventilation, and death [3–6]. Individuals from some ancestry groups such as Native American, African American or Black, and Hispanic individuals appear to be at higher risk of both SARS-CoV-2 infection and severe COVID-19 [7]. Individuals with underlying medical conditions: chronic kidney disease [5], chronic obstructive pulmonary disease, immunocompromised conditions, hypertension, and Type 2 diabetes mellitus have been reported as having higher risk for severe illness from COVID-19 [3,6,8–13]. Other risk factors continue to emerge, but an unprecedented need remains for research to further understand COVID-19 risk factors and the impact of shelter in place or quarantine on long-term risks for other diseases.

We developed the Michigan Medicine Precision Health COVID-19 Survey to evaluate SARS-CoV-2 exposure, COVID-19 symptoms and risk factors, and the impact of the ‘Stay Home Stay Safe’ Executive Order on previously enrolled Michigan Medicine biorepository participants. We aimed to answer three main questions with this study. First, which risk factors are associated with contracting COVID-19, and are they different from risk factors that are associated with a severe COVID-19 course? Secondly, why are African-Americans at higher risk of COVID-19? And lastly, is there a potential impact of the ‘Stay Home Stay Safe’ executive order in Michigan on other health behaviors that may relate to the risk of developing long-term cardiometabolic disease?

## METHODS

Prior to March 2020, Michigan Medicine biorepository participants provided broad consent for biospecimen collection, electronic health data, future and ongoing use of data for undefined research, and re-contact in future studies [14]. On March 12, Michigan’s Governor Gretchen Whitmer ordered all schools to close after the first two confirmed cases of COVID-19 in the state were reported on March 10. Then, on March 23, Governor Whitmer issued the ‘Stay Home Stay Safe’ Executive Order, which enforced school closures, ordered non-essential workers to work from home, enforced restaurant and bar closures (with the exception of contactless take-out), and discouraged socializing, travel and unnecessary trips outside of the house. SARS-CoV-2 testing was limited, and typically only available to symptomatic individuals or frontline healthcare workers during the time prior to survey deployment.

We developed the COVID-19 survey to evaluate risk factors for COVID-19 and the impact of the ‘Stay Home Stay Safe’ executive order on health behaviors of biorepository participants, primarily from the Michigan Genomics Initiative (MGI, 74,194 enrolled to date) [15], the Cardiovascular Health Improvement Project (CHIP, 5,708 enrolled to date) [16,17], and the Michigan study of Racial Equality and Community Health (MREACH, ~300 enrolled to date, 85% of whom are African-American or Black) (Supplemental Table 1, description of all biorepository cohorts). Of the >75,000 Michigan Medicine biorepository participants, 50,512 participants had a valid email address in their electronic health records. Up to 3 survey invitations were sent to each participant by email between May 26, 2020 and June 29, 2020, which was 10-15 weeks after schools were closed. Survey responses received prior to July 2, 2020 were included in analysis. Daily positive COVID-19 tests for the state of Michigan relative to the time of survey deployment are presented in Figure 1 [18].

**Figure 1.**
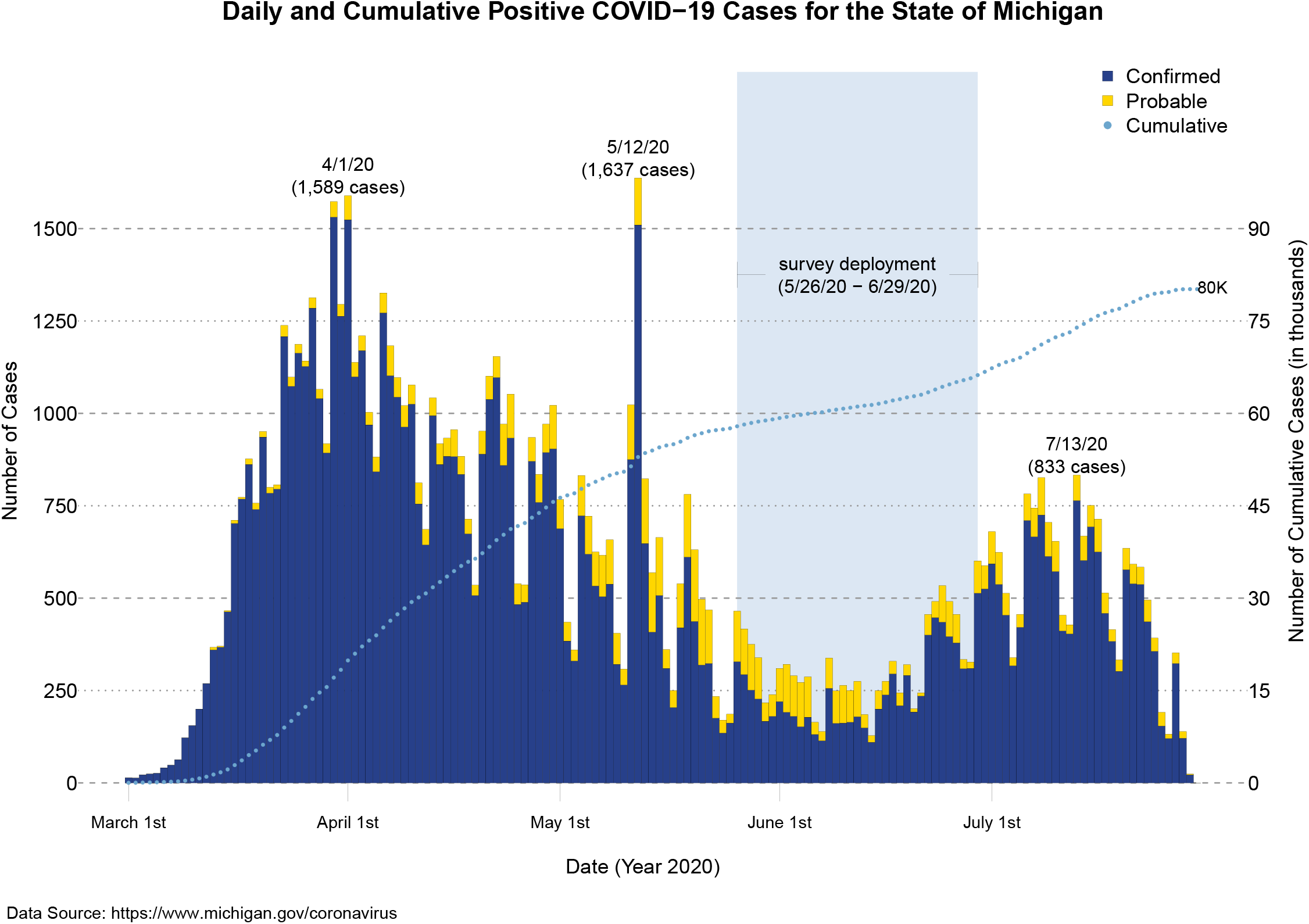
Confirmed and probable COVID-19 cases for the state of Michigan from March 1, 2020 to July 29, 2020.

Of 50,512 participants with valid email addresses, 8,422 (17%) completed the survey. 375 survey respondents (4.5%) were excluded from analysis for the following reasons: international location/zip code (n=13), sex discrepancy between electronic health record and survey response (n=236), and self-diagnosis of COVID-19 without test (n=134). We were not able to confirm COVID-19 self-diagnoses and therefore, these were excluded (Supplemental Table 2 provides descriptive statistics for these participants). A total of 8,047 survey respondents were included in analysis, 133 of whom were diagnosed with COVID-19 by a doctor or test (Figure 2). European-Americans were slightly more likely to complete the survey (17.3% of those invited) relative to African-Americans (15.4% of those invited; Supplemental Table 3).

**Figure 2.**
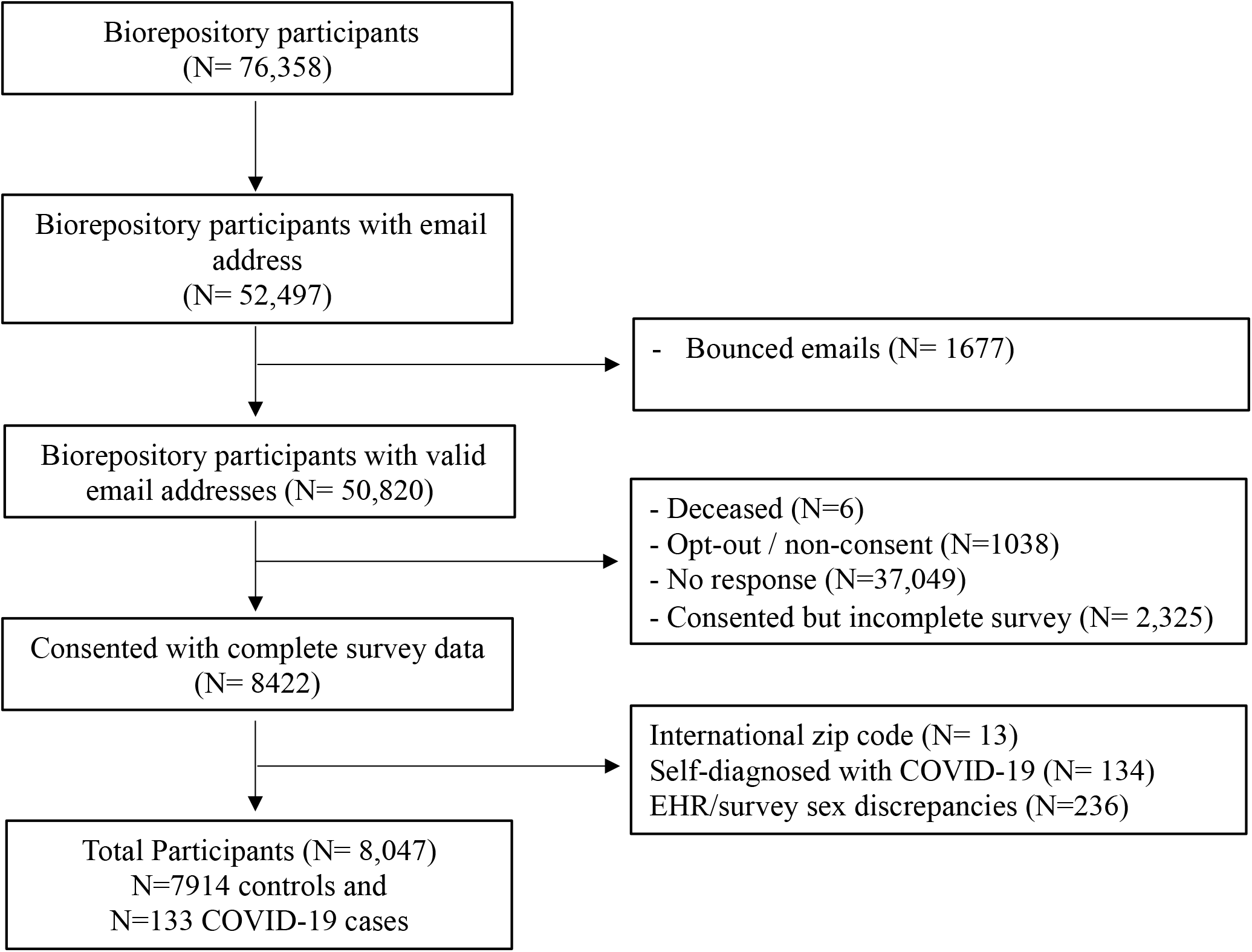
Study Enrollment.

### Survey Deployment

The University of Michigan Data Office for Clinical and Translational Research (DOCTR) deployed the survey to biorepository participants’ email addresses by sending an individualized link to a Qualtrics electronic informed consent and survey. The DOCTR office maintained personal health information and created coded identifiers for researchers to access survey data [19]. The protocol and study procedures were approved by the University of Michigan Institutional Review Board (HUM00180827).

### Survey Content

The survey evaluated self-reported SARS-CoV-2 exposure, COVID-19 diagnosis, symptoms, and risk factors as well as health behaviors and overall concern during the Michigan ‘Stay Home Stay Safe’ Order. Skip logic was applied, meaning most participants only answered 50% of the 96 total questions, requiring approximately 8-10 minutes. A full version of the survey with branching logic is available (Supplemental Table 4).

#### Participant Characteristics

The survey included basic demographics including sex, race/ethnicity, which were used for analysis. Electronic health records were utilized to verify self-reported sex, as well as demographics of survey non-respondents (Supplemental Table 3). Participants were asked to report height and weight for calculation of body mass index. Socioeconomic status was assessed by self-reported average annual household income, which was categorized as low (<$40,000), middle ($40,000 to $100,000), or high (>$100,000).[20]. Socioeconomic indicators of health were evaluated, including current living situation (e.g., owned versus rented accommodations), zip code for ‘Stay Home Stay Safe’ shelter in place location, highest grade or year of school completed, cars/automobiles per household, primary mode of transportation[21].

#### COVID-19 Exposure, Symptoms, Diagnosis, and Severity

All participants were asked whether they were diagnosed with COVID-19 with a test or by a physician without a test [22]. All participants responding “no” to being diagnosed with COVID-19 were classified as controls, with the exception of participants who thought they might have or have had COVID-19 but had not been tested or diagnosed by a physician. Possible self-diagnosed individuals were excluded from all standard analyses (Supplemental Table 2, symptoms are reported). All participants were asked to recall potential COVID-19 exposure, including international and domestic travel history, current work as or living with an essential employee, and contact with family members or someone outside the house diagnosed with COVID-19. COVID-19 cases were asked to recall symptoms, duration of symptoms, hospitalization, and complications. Participants reporting symptoms and shortness of breath without hospitalization were categorized as mild-to-moderate whereas participants hospitalized due to COVID-19 were categorized as severe [23].

#### Medical and Family History

Participants were asked to recall history of chronic diseases including respiratory, immune, genitourinary/metabolic, cardiovascular, neurological, and oncological conditions.

#### Mental Health

Participants were asked to recall their level of concern about the COVID-19 pandemic and their concerns for contracting the virus, financial situation, and isolation. Participants were asked to recall personal precautions, where they obtained COVID-19, and whether family members have been diagnosed with or died from COVID-19.

#### Health Behaviors

Participants were asked to recall how health behaviors may have changed during the ‘Stay Home Stay Safe’ period relative to usual behavior. Specifically, participants were asked to recall whether they had increased moderate-to-strenuous exercise, alcohol consumption, drug use, and tobacco use or whether they had improved sleep habits and nutrition, and whether or not they had gained weight. These items were assessed using a Likert 5-point scale (Strongly Disagree to Strongly Agree). Participants were asked to recall for an average week (before and after COVID-19 ‘Stay Home Stay Safe’ recommendations), how many days they participated in a total of 30 minutes or more of physical activity[24]. Information related to smoking history was assessed.

### Statistical Analysis

The risk factors studied were demographic variables, socioeconomic indicators, environmental factors, health behaviors, workplace information, community exposure, precautions practiced, and comorbidities collected from the survey. P-values < 0.05 were applied to define statistical significance.

To test for the association between risk factors and COVID-19 status, we performed logistic regression. When assessing the association with clinical risk factors, such as type 2 diabetes, we included age and sex as covariates. We further categorized disease status into mild-to-moderate and severe patients (requiring hospitalization) as a secondary outcome to test for clinical and social risk factors associated with disease severity. To evaluate the effects of ethnicity (African American versus European American) on COVID risk factors, a chi-square test or t-test were applied for categorical and continuous risk factors, respectively. Lastly, to study the impact of the ‘Stay Home Stay Safe’ executive order on participants’ behavioral changes and level of concern -- differences by ethnicity, sex, and income groups were evaluated. Ordinal logistic regression was used to examine the association between ordinal behavioral change (disagree, same, and agree) as the dependent variable and ethnicity, sex, and income group as independent variable. Logistic regression was used to examine the association between binary outcomes of level of concern. Linear regression was applied to evaluate association between a continuous scale of concern level (range 1-10 where 10 is a high level of concern) and ethnicity, sex, and income group as the independent variable.

For health behavioral change questions, participants were given five categories in the answer field. We collapsed “strongly disagree” and “disagree” into one group, kept “about the same”, and collapsed “strongly agree” and “agree” into another group as a three-group ordinal outcome to improve statistical power. Health behavior change on tobacco use was only evaluated on participants who are current users or switched tobacco/nicotine products from one to another, and drug use behavior change was only evaluated on individuals who answered they have used opioid, benzodiazepines, or marijuana/cannabis in the past 30 days. Level of concern categorical responses (not concerned, slightly concerned, very concerned, and extremely concerned) were dichotomized into “No” (not concerned and slightly concerned) and “Yes” (very concerned and extremely concerned) as a binary outcome.

## RESULTS

Of 50,512 biorepository participants invited to participate in a COVID-19 survey, a total of 8,047 participants are included in this cross-sectional analysis (Supplemental Table 3) and 133 (1.7%) participants responding “yes” to being diagnosed with COVID-19 by a test or a physician. Among all survey respondents, the mean age was 59 ± 15 (mean ± SD) years old and 3,314 (41.2%) were male. Among survey respondents, 234 (2.9%) self-identified as Black or African American (hereafter referred to as African American), 7,389 (91.8%) as White/Caucasian (hereafter referred to as European American), and 424 (5.3%) as another category (American Indian or Alaska Native, Asian, Native Hawaiian or Pacific Islander, Unknown, or Prefer not to answer).

Participants reported high socioeconomic status, with more than forty-two percent reporting an annual household income greater than $100,000, and 67% earned a bachelor’s degree or higher. The average BMI was 29 ± 7 with 33.5% and 37.4% categorized as overweight and obese, respectively. The University of Michigan / Michigan Medicine is in Ann Arbor, which has higher rates of education and salary than national norms. However, based on our data of COVID-19 related symptoms start date, we observed a similar pattern between number of patients started their symptoms and number of confirmed cases in Michigan, which indicated the high generalizability of this dataset (Figure 3).

**Figure 3.**
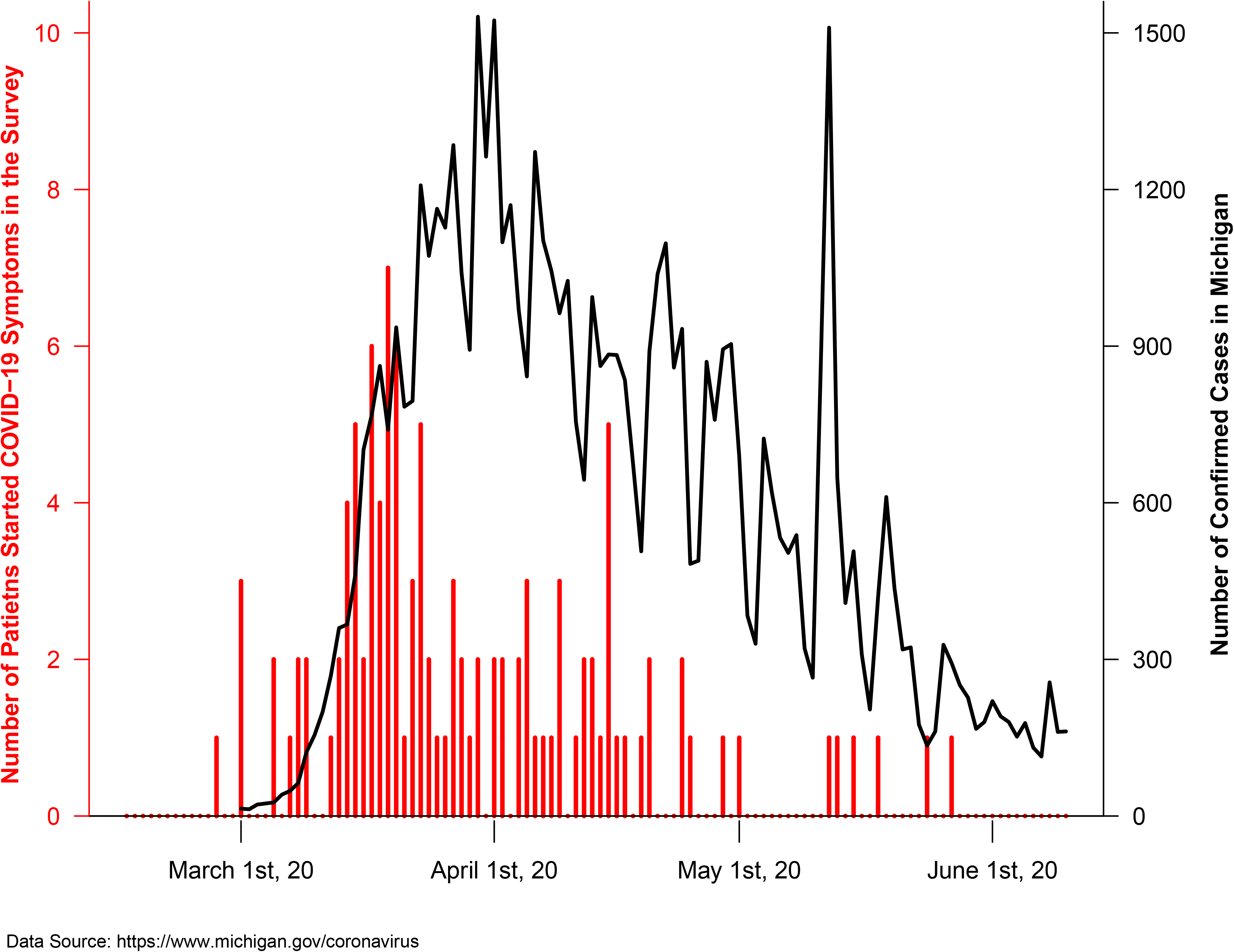
Number of patients by symptoms start date in the survey and number of confirmed cases in Michigan.

### Risk factors associated with developing COVID-19

We examined demographic differences between COVID-19 confirmed cases and controls. Participants who self-reported as “African-American or Black” were significantly more likely to be diagnosed with COVID-19 compared to self-reported “White/Caucasian” individuals (5.6% versus 1.5%, p=1×10^-4^). This approximate 3-fold higher risk of developing COVID-19 we observed in African Americans is consistent with Washtenaw County demographics of COVID-19 cases (as of July 16, 2020) where 32% of COVID-19 lab-confirmed cases are Black or African-American whereas only 12.3% of the Washtenaw County population is Black or African-American (Supplemental Table 5).

The data showed that people with any of the COVID-19 symptoms collected from the survey were at a significantly higher risk of contracting COVID-19. More than half of the COVID-19 cases reported having fatigue, muscle aches, shortness of breath, headache, cough, or fever (ordered by most common to less common). (Supplemental Table 2 for self-diagnosed).

Next, we evaluated possible sources of exposure to SARS-CoV2 among the cases (Figure 4a, Supplemental Table 6). COVID-19 cases were younger (51±15 versus 59±15 years, p=6×10^-10^), had more social exposure to others with COVID-19 (family members [33.0% versus 6.8%, p=7×10^-8^] and people outside of household [20.9% versus 9.9%, p=0.046]) than controls. COVID-19 cases were more likely to report their role as an essential employee (45.1% versus 19.4%, p=4×10^-12^) and medical professional (24.8% versus 8.0%, p=8×10^-11^). Avoiding public transport (65.4% versus 76.2%, p=5×10^-3^) and self-isolation (17.3% versus 26.6%, p=0.012) were associated with significantly lower risk of contracting COVID-19, but most of the personal precautions were not found to be individually associated with reduced COVID-19 risk. However, not doing any of the precautions significantly increased the risk of contracting COVID-19 (2.3% versus 0.3%, p=0.002). Several precautions were reported to be used at high rates amongst all survey respondents (e.g. 95% report mask wearing, 96% report frequent hand-washing, and 92% report social distancing); therefore, there was little power to distinguish any difference in infection due to not taking these precautions. Despite high rates of self-reported personal precautions, 58.3% of COVID-19 cases reported no known exposure to a COVID-19-positive individual in the two weeks prior to their diagnosis. Using age and sex as covariates, clinical risk factors significantly associated with COVID-19 were: type 2 diabetes (15% versus 11.8%, p=0.022), respiratory conditions (42.9% versus 36%, p=0.025), and congestive heart failure (6% versus 3.7%, p=0.035).

**Figure 4.**
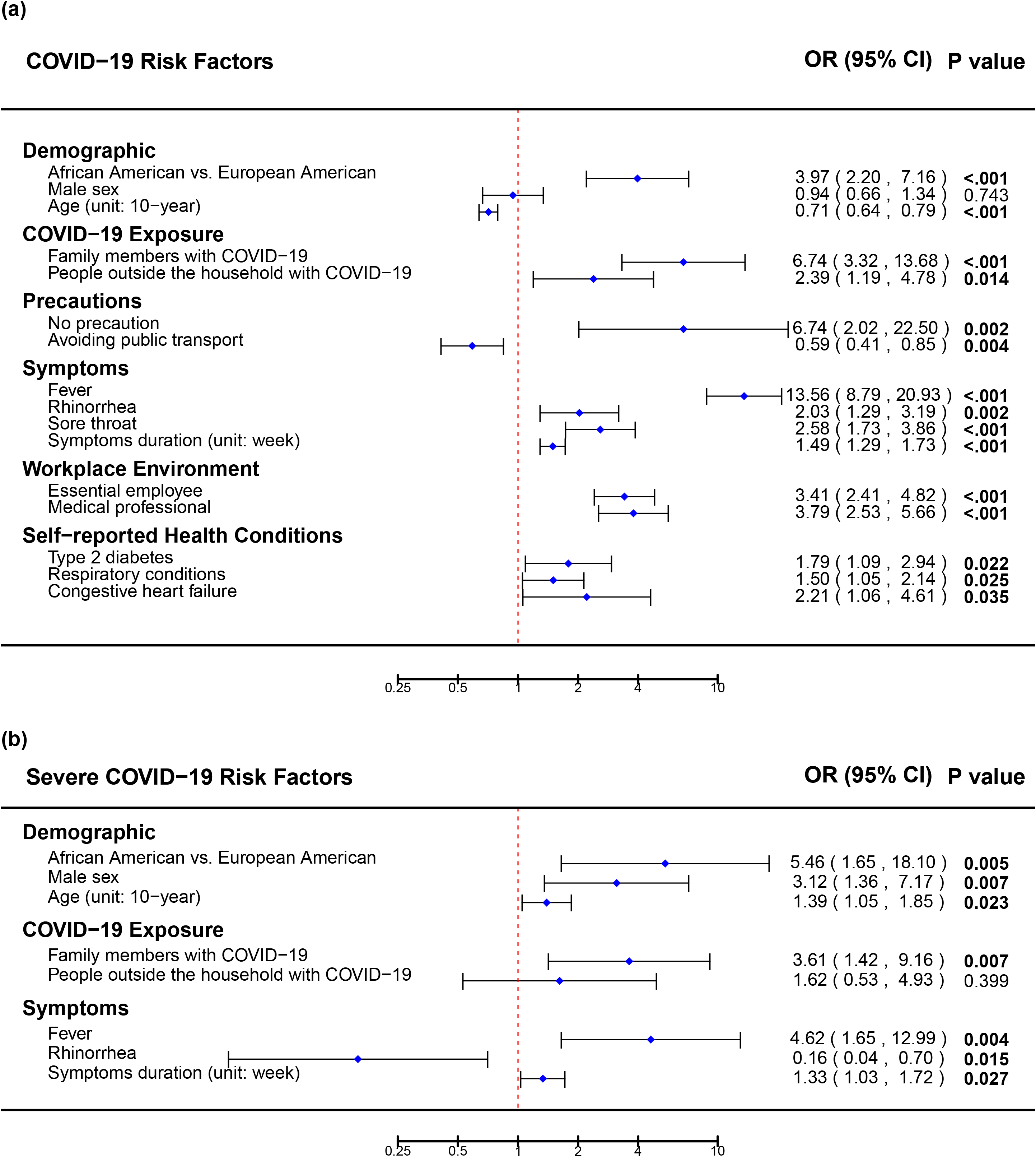
**a)** Risk factors associated with COVID-19 in comparison to those without COVID-19, and **b)** risk factors associated with a severe course of COVID-19 in comparison to those with a mild or moderate course of COVID-19.

### Risk factors associated with a severe COVID-19 disease course

Next, factors associated with a severe course of COVID-19 were examined (Figure 4b, Supplemental Table 6). We classified 31 cases as severe based on being hospitalized for COVID-19 compared to 102 mild-to-moderate cases who did not require hospitalization. African Americans were more likely to have had a severe COVID-19 disease course compared to European American (53.8% versus 17.6%, p=0.005), however this sample size was limited. Severe cases were more likely to be male (61.3% versus 38.7%, p=0.007), older (57±14 versus 49±15 years, p=0.023), and report social exposure to COVID-19 (family members diagnosed with COVID-19 (53.6% versus 24.2%, p=0.012). Severe cases reported that COVID-19 symptoms persisted an average of 23±11 days compared to 17±11 days for mild-to-moderate cases (p=0.027). A higher proportion of patients with severe COVID-19 reported fever (83.9% versus 52.9%, p=0.001). Conversely, mild-to-moderate patients were more likely to report rhinorrhea (30.4% versus 6.5%, p=0.003) compared to severe patients.

### Possible explanations of why African-Americans at higher risk of COVID-19

To attempt to understand why African Americans were at higher risk of COVID-19, we evaluated socioeconomic status, COVID-19 exposure, and environmental factors by ethnicity in the entire set of survey respondents (Figure 5, Supplemental Table 7). African American survey respondents were younger (53±15 versus 60±15 years, p=1×10^-9^), more likely to be female (69.5% versus 58.3%, p=8×10^-4^), have a higher BMI (32.5±8.1 versus 29.0±6.6, p=9×10^-10^), and report higher rates of obesity (BMI of 30.0 or higher, 57.8% versus 36.9%, p=3×10^-9^). African Americans reported a lower income (annual family income < $40,000; 28.1% versus 13.4%, p=1×10^-9^), higher rates of living in rental housing (31.6% versus 9.1%, p=5×10^-30^), and more social exposure to COVID-19-positive individuals (family members [39.1% versus 13.5%, p=0.005]; people outside of the household with COVID-19 [38.9% versus 11.0%, p=0.003]). African Americans were more likely to report being an essential employee during ‘Stay Home Stay Safe’ (26.6% versus 19.4%, p=0.008), including being a medical professional (13.7% and 8.0%, p=0.002). Self-reported precautions taken to avoid COVID-19 were not different between African and European Americans in this survey and could not explain the difference in rates of COVID-19. In fact, we observed (non-significantly) higher rates of precautions in African Americans than European Americans.

**Figure 5.**
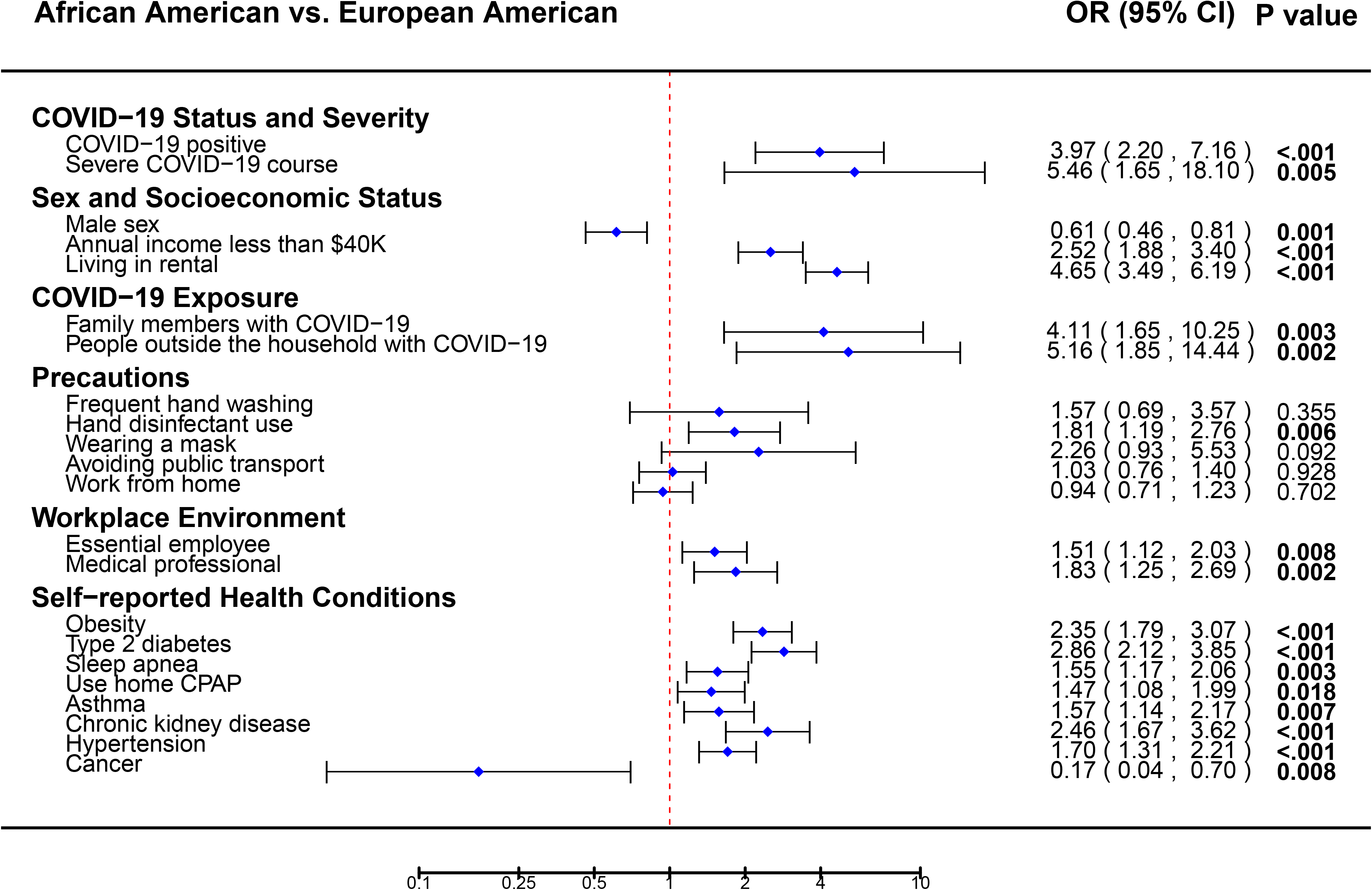
Differences in clinical and social risk factors between African American and European American survey respondents.

African Americans were more likely to suffer from a severe COVID-19 response (53.8% hospitalized versus 17.6% for European-American, p=0.005) in this dataset, but the sample size of hospitalized patients was small (N=31). We next evaluated self-reported disease risk factors by ethnicity that may increase risk of a severe course of COVID-19, or other cardiometabolic diseases (Figure 5, Supplemental Table 7). African American survey respondents reported a significantly higher (p-value < 0.05) incidence of: type II diabetes mellitus (26.5% versus 11.2%), sleep apnea (30.3% versus 21.9%), use of CPAP (23.5% versus 17.3%), asthma (20.9% versus 14.4%), chronic kidney disease (13.7% versus 6.0%), hypertension (44.9% versus 32.4%), and obesity (57.8% versus 36.9%), while European Americans reported a higher incidence of cancer (4.8% versus 0.9%).

### Health behaviors during statewide ‘Stay Home Stay Safe’ order

We also examined the impact of the ‘Stay Home Stay Safe’ period in Michigan on health behaviors, lifestyle changes, and level of concern of survey participants. We examined if there were differences in health behaviors within groups divided by sex, by three income groups, and by ethnicity (Figure 6, Supplemental Table 8, 9, and 10). We excluded the 133 individuals diagnosed with COVID-19 because their lifestyle may have been greatly impacted by the disease itself. This comparison was meant to help us understand the potential increase in risks for cardiometabolic or other diseases imposed by the ‘Stay Home Stay Safe’ period and to determine if the restrictions impacted some groups more than others.

**Figure 6.**
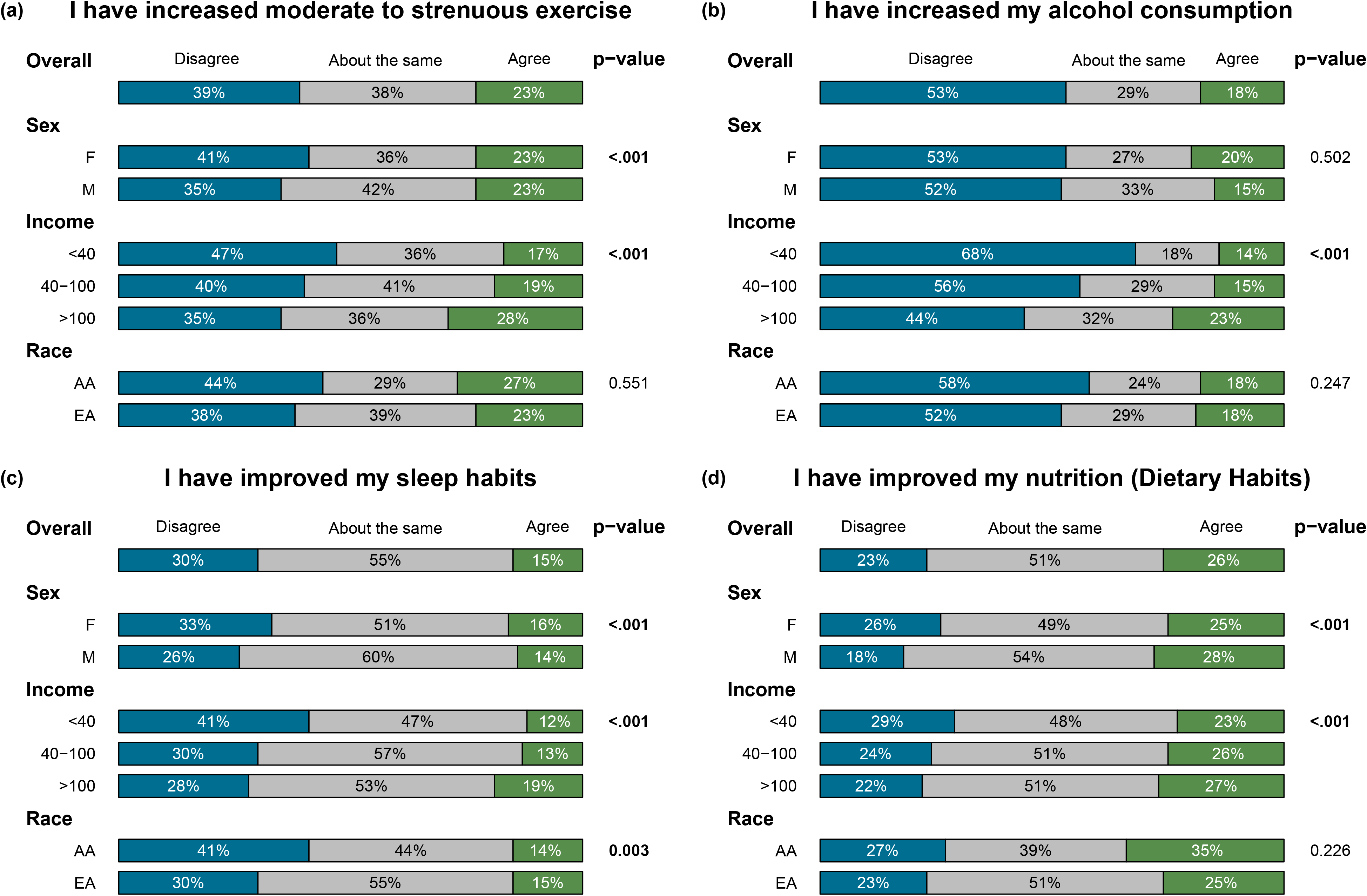
Behavioral change by sex (Female versus Male), income (<$40,000 versus $40,000-100,00 versus >$100,000 annual house income), and race (African versus European American).

First, across all participants without COVID-19, we found that a reasonable proportion of participants developed less-healthy behaviors during the ‘Stay Home Stay Safe’ period: 23.1% report worsened nutrition, 31.9% report weight gain, 30.2% report poorer sleep habits, 38.6% report decreased moderate-to-vigorous exercise, 18.2% report increased alcohol consumption, 35.9% of current smokers report increased smoking, and 12.7% of current drug users report increased drug use. On the other hand, a substantial proportion reported healthier behaviors: 26.0% reported better nutrition, 32.4% reported weight loss, 15.2% reported improved sleep habits, 23.0% reported increased moderate-to-vigorous exercise, 52.7% reported decreased alcohol consumption, 28.2% reported decreased smoking, and among drug users, 68.3% reported decreased drug use.

During ‘Stay Home Stay Safe’, when compared to men, women were significantly more likely to report behavioral changes that could increase the risk of cardiometabolic diseases: worsened nutrition (26.4% versus 18.3%, p=4×10^-12^), weight gain (36.6% versus 25.1%, p=6×10^-21^), poorer sleep habits (33.2% versus 25.7%, p=1×10^-5^), increased tobacco use among tobacco users (43.7% versus 25.3%, p=6×10^-4^), and decreased moderate-to-vigorous exercise (41.1% versus 34.8%, p=2×10^-4^). Conversely, men were significantly more likely to report improved nutrition and weight maintenance. The majority of the men reported that they kept their exercise, sleep habits, and tobacco use the same during the ‘Stay Home Stay Safe’ executive order (Figure 6, Supplemental Table 8).

We also examined the association between household income and health behavior changes during ‘Stay Home Stay Safe’ (Figure 6, Supplemental Table 9). People with higher income were more likely to report increased exercise (28.3% versus 19.4% versus 17.1%, p=2×10^-17^), increased alcohol consumption (23.4% versus 15.1% versus 13.5%, p=5×10^-38^), and also improved sleep habits (19.1% versus 12.8% versus 12.0%, p=8×10^-17^) and nutrition (27.4% versus 25.8% versus 23.3%, p=6×10^-5^). People with higher income were also more likely to report working from home during ‘Stay Home Stay Safe’ (53.5% versus 27.9% versus 17.1%, p=2×10^-146^). People with lower income were more likely to report weight gain (36.7% versus 31.5% versus 31.7%, p=0.029).

Of the behavioral changes made, there were few statistically significant differences by ethnic group (Figure 6, Supplemental Table 7 and 10). Of all behavioral categories surveyed, we only observed a significant difference for exercise between African Americans relative to European Americans. African Americans reported less exercise per week (days of 30 minutes or more of physical activity in an average week) from both before (3.3±1.8 versus 3.7±1.9 days, p=0.003) and after the COVID-19 pandemic (3.1± 1.9 and 3.8± 2.1 days, p=6×10^-5^). African Americans were also more likely to report decreased exercise during ‘Stay Home Stay Safe’ than European-Americans (31.3% versus 24.2%, p=0.020).

When asked about overall concern (range 1-10 where 10 is a high level of concern), the population mean was 5.57±2.95. When asked about specific aspects, 47.4% reported concern about contracting COVID-19, 62.3% had concern of someone close to them contracting COVID-19, 18.3% had concern about serious financial problems, 10.6% were concerned about losing their job, 51.4% were concerned that it will be a long time before life returns to normal, and 53.6% were concerned about not seeing friends and family.

Women report higher levels of overall concern than men (5.69±2.88 versus 5.39±3.05 p=6×10^-6^), and more women report concern about people close to them contracting COVID-19 (64.4% versus 59.4%, p=7×10^-6^), developing serious financial trouble (20.3% versus 15.4%, p=3×10^-8^), losing their job (12.4% versus 8.1%, p=2×10^-9^), concern that it will be a long time before life returns to normal (54.8% versus 46.6%, p=7×10^-13^), and not seeing friends and family (56.8% versus 49.1%, p=2×10^-11^) (Supplemental Table 8).

Relative to the high income group (>100K), people with lower or medium income (<40K or 40K-100K) had higher levels of concern about (Supplemental Table 9): contracting COVID-19 (49.3% versus 48.4% versus 44.9%, p=0.006), getting into serious financial trouble (34.8% versus 20.4% versus 11.8%, p=7×10^-59^), losing their job (13.7% versus 11.4% versus 9.3%, p=1×10^-4^), and that it will be a long time before life returns to normal (55.1% versus 51.3% versus 50.6%, p=0.035). When we examined groups by self-reported ethnicity (Supplemental Table 10), African Americans report higher overall concern (6.72±3.01 versus 5.53±2.94 p=3×10^-9^) as well as concerns about: contracting COVID-19 (63.3% versus 47.0%, p=3×10^-6^), people close to them contracting COVID-19 (71.5% versus 62.1%, p=0.005), developing serious financial problems (33.9% versus 17.5%, p=1×10^-9^), losing their job (20.4% versus 10.1%, p=2×10^-6^), and concern that it will be a long time before life returns to normal (58.4% versus 51.3%, p=0.039).

## DISCUSSION

In this study, we first sought to determine which demographic, clinical or behavioral risk factors might predispose individuals to COVID-19, and determine if the same or different risk factors might predispose to a severe COVID-19 disease course. Based on this survey of 133 COVID-19 cases and 7,914 controls, we were able to identify significant risks of COVID-19 for individuals who are: African American, younger age, essential employees and those who report being exposed to other COVID-19 cases including family and others outside the household. The most common symptoms among cases were fatigue (78.9%), muscle aches (66.9%), and shortness of breath (65.4%), whereas sneezing (16.5%) and runny nose (24.8%) were less common. Personal precautions against transmission appeared to decrease spread of SARS-CoV2, and individuals who reported using no precautions were at higher risk of COVID-19.

African Americans account for 14% of the state of Michigan’s population, but 33% of COVID-19 cases and 41% of deaths (data as of July 16, 2020), which is consistent with observations at the national level [25]. Given the alarming disparity, we examined potential risk factors that may explain the higher rates of COVID-19 observed among African American individuals. Our data identified a number of clinical and social risk factors that were different between African American and European American participants. Several chronic diseases (obesity, hypertension, type II diabetes, and chronic kidney disease) had higher rates in African Americans by self-report. Based on significant differences between African Americans and European Americans in annual income, designated essential employees, and different rates of living in rented accommodation, we hypothesize that African Americans were at a higher risk of contracting COVID-19 because of economic pressure to continue working and interacting with people outside the household during the ‘Stay Home Stay Safe’ order in Michigan (Figure 7). Larger studies are needed to tease apart which risk factors are explicitly driving the higher incidence rates of COVID-19 in African Americans.

**Figure 7.**
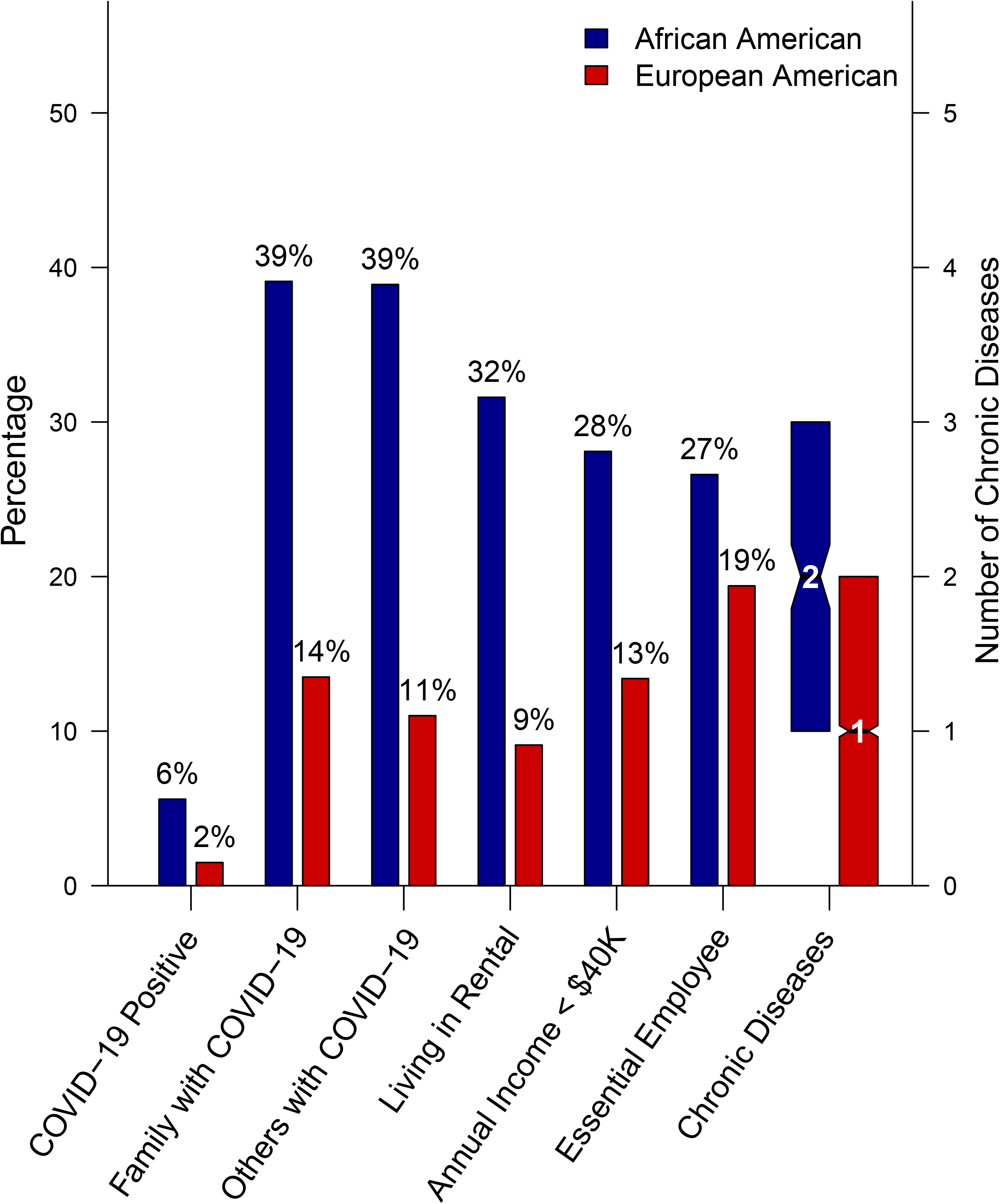
COVID-19 status, COVID-19 risk factors, and chronic disease differences by race.

The long-term effects of the pandemic may further exacerbate the higher incidence and severity of chronic and cardiometabolic diseases in some demographic groups. Given this possibility, we evaluated the impact of COVID-19 on self-reported health behaviors. Interestingly, the impact on health behaviors were variable, but on average, we found that men were more likely to stay the same or improve health behaviors such as, exercise, sleep habits, tobacco use, and nutrition. Conversely, women were more likely than men to report less-healthy behaviors, including worsened nutrition, weight gain, poorer sleep habits, and decreased exercise. Women and African Americans report higher levels of overall concern, concern about people close to them contracting COVID, developing serious financial trouble, losing their job, concern that it will be a long time before life returns to normal, and not seeing friends and family. People with higher income were more likely to increase exercise, increase alcohol consumption, and also improve sleep habits and nutrition. People with lower income were more likely to gain weight, also had higher levels of concern about contracting COVID-19, getting into serious financial trouble, losing their job, and that it will be a long time before life returns to normal.

This study has several limitations that should be considered when reviewing the findings. First, this study was conducted at a single center with a limited geographic area and it utilized a retrospective design. The study design engaged previously consented biorepository participants to enroll and recall their COVID-19 exposures, symptoms, diagnosis, precautions, and experience. It is possible that there is respondent bias as participants from the various biorepositories may have responded differently. Furthermore, we excluded 134 individuals who were self-diagnosed with possible COVID-19 but were not diagnosed by a doctor or a test. We cannot determine the impact of personal precautions reported to be used at high rates, such as mask wearing and frequent hand washing because of lack of power. Additionally, we do not have power to distinguish which socioeconomic, employment exposure or health factors that differ between ethnic groups may be the cause of higher COVID-19 rates in African-American. Lastly, because our survey was only taken once, we did not capture longitudinal data, or information on pre-symptomatic individuals who later tested positive.

Understanding exposure risks are critical to educating the public and to saving lives. Our data provides insight into exposure risks, confirms that precautions work, although being an essential worker or medical professional increases the susceptibility of transmission. Overall, African Americans, women, and those with low household income reported less healthy behaviors during the ‘ Stay Home Stay Safe’ (post-COVID) period in Michigan, while also reporting more overall concern for possible economic, health and societal decline related to the global COVID-19 pandemic. There is an undeniable need to focus continued efforts on prevention and mitigation strategies for COVID-19, and begin to more comprehensively address the inequality gaps in disease risks by ethnic group that the COVID-19 pandemic has highlighted.

## Data Availability

Data is available from the Data Office at the University of Michigan for IRB approved investigators.

## ACKNOWLEDGEMENTS

The authors acknowledge the participants, recruitment team and project managers of the Michigan Genomics Initiative, Cardiovascular Health Improvement Project, Michigan study of Racial Equality and Community Health, the University of Michigan Medical School Research Data Warehouse and Precision Health Initiative resources for providing data aggregation, management, and distribution services in support of the research reported in this publication. We acknowledge the University of Michigan Rogel Cancer Center and the Michigan Institute of Data Science.

## Notes

### Competing Interest Statement

The spouse of Dr. Willer works for Regeneron Pharmaceuticals Inc. Dr. Brummett is a consultant for Heron Therapeutics and Alosa Health -- not related to the present study.

### Funding Statement

Dr. Willer is supported by National Institutes of Health grants R35-HL135824, R01-HL142023, R01-DK075787, and R01-HL109946
Dr. Douville is supported by the Foundation for Anesthesia Education and Research (FAER) - Mentored Research Training Grant.

### Author Declarations

University of Michigan MED-IRB (HUM00180827)

